# Should healthcare workers with SARS-CoV-2 household exposures work? A Cohort Study

**DOI:** 10.1101/2022.01.23.22269719

**Authors:** Caroline Quach, Ana C. Blanchard, Josée Lamarche, Nathalie Audy, Valérie Lamarre

**Author notes:** Corresponding author: Caroline Quach., CHU Sainte-Justine, 3175 Cote Ste-Catherine, suite 7.17.008, Montreal (QC) Canada H3T 1C5.

## Abstract

**Importance:** Due to high community transmission of the Omicron variant, healthcare workers (HCWs) have been increasingly reporting household exposures to confirmed COVID-19 cases. Quebec (Canada) provincial guidelines required to quarantine these HCWs. Facing the risk of staffing shortages, our hospital decided to allow them to work.

**Objective:** To evaluate the risk for HCWs, who were household contacts, to become positive for COVID-19 by RT-PCR and evaluate the risk of nosocomial COVID-19 transmission.

**Design:** Cohort of HCWs with a history of household exposure to a confirmed case of COVID-19.

**Setting:** CHU Sainte-Justine, a tertiary care mother and child center in Montreal (QC) Canada

**Participants:** Consecutive HCWs who contacted OHS between December 20, 2021 and January 17, 2022 for a history of household exposure to COVID-19.

**Exposure:** Confirmed case of COVID-19 in the household

**Main outcome and measures:** The main outcome was a positive RT-PCR for SARS-CoV-2. Outbreaks and nosocomial cases were identified through daily analysis of COVID-19 cases, by sector and part of the usual Infection Prevention and Control surveillance process.

**Results:** Overall, 237 of 475 (50%) HCWs who declared a known household contact with a confirmed COVID-19 case remained negative. Of those who became positive, 196 (82.4%) were positive upon initial testing and were quarantined. Only 42 (15%) of 279 HCWs who were allowed to work became positive, a median of 4 days after the initial test. The absence of symptoms at initial evaluation (OR 3.8, 95% CI 2.5-5.7) and having received a third vaccine dose more than 7 days before (OR 1.88, 95% CI 1.3 – 2.8) were associated with an increased odds of remaining negative. There was no outbreak among HCWs and no nosocomial transmission to patients from a HCW that was allowed to work, while a known household contact.

**Conclusion and relevance:** Measures taken to protect the health care environment from COVID-19 must be cautiously balanced with the risk of staffing shortage. Allowing vaccinated asymptomatic HCWs who are known household contacts of confirmed COVID-19 cases to work is likely a safe alternative, when staff shortage is anticipated.

## Introduction

In the province of Quebec (Canada), the Omicron variant of concern (VOC) became predominant as of December 20 ^th^, 2021 (1). Due to high community transmission, healthcare workers (HCWs) were increasingly reporting significant household exposures to confirmed cases. Previous provincial recommendations allowed HCWs to work despite contact with a confirmed SARS-CoV-2 infected household member if HCWs were: 1) vaccinated with 2 doses of COVID-19 vaccine > 7 days before contact and 2) asymptomatic (2). With emergent data showing a low 2-dose effectiveness against Omicron, increasing from 30-40% to 75% after a booster dose (3), new provincial guidelines (4) considered everyone as unprotected, quarantining all household contacts and isolating infected HCWs for 10 days, in most circumstances.

Facing the risk of staffing shortages, our hospital decided to allow HCWs to work, despite significant household exposures. These HCWs had to have received ≥2 doses of vaccine and follow exemplary measures (procedure masks/N95 permanently, eating alone in a closed room, symptoms monitoring, and reverse-transcription polymerase chain reaction (RT-PCR) for SARS-CoV-2 every 3 days until 7 days after the end of the index case’s period of contagion). The objective of this study was to evaluate the risk for HCWs, who were household contacts, to become positive for COVID-19 by RT-PCR and evaluate the risk of nosocomial COVID-19 transmission.

## Methods

### Study Design

Starting on December 20^th^, 2021, we followed a cohort of HCWs who contacted the Occupational Health and Safety (OHS) unit of CHU Sainte-Justine through the established process, to report a household contact with a confirmed COVID-19 case.

### Setting and participants

CHU Sainte-Justine is a tertiary care mother and child hospital located in Montreal (QC), Canada. Since the beginning of the COVID-19 pandemic, OHS established a call center, staffed 24/7 with nurses who evaluate all HCWs with symptoms or exposures, under the supervision of Human Resources and the medical direction of the Infection Prevention and Control (IPAC) physicians. A testing clinic is also available on-site for RT-PCR, which are done in our diagnostic microbiology laboratory, as previously described (5).

### Variables and measurement

The main outcome was a positive RT-PCR. Collected variables included vaccine (doses/dates), RT-PCR (results/dates), self-reported symptoms at initial test, cases of nosocomial COVID-19 cases in patients, and outbreaks among HCWs. Outbreaks and nosocomial cases were identified through daily analysis of COVID-19 cases and reported to the Quebec Ministry of Health and Social Services, as part of the usual IPAC surveillance process. A case was suspected to be nosocomial if symptoms onset occurred ≥3 days after admission.

### Statistical analysis

We used descriptive statistics for the proportion of HCWs who became RT-PCR positive and used a Kaplan-Meier curve, stratified on symptoms at baseline and on 3 ^rd^ dose vaccination status (valid if > 7 days). We used logistic regression to determine variables associated with the risk of a positive RT-PCR. All analyses were done using STATA version 17.0 (StataCorp LLC, College Station, TX).

### Ethical consideration

Because this was an quality improvement evaluation using data collected through our usual process of care, we obtained a waiver from the Research Ethics Committee.

## Results

From December 20^th^, 2021 to January 17^th^, 2022, a total of 929 HCWs were positive for COVID-19 by RT-PCR. During the same period, 475 HCWs declared a known household contact with a confirmed COVID-19 case, 237 (49.9%) remained negative. Table 1 summarizes baseline characteristics. Of those who were positive by RT-PCR, 196/238 (82.4%) were positive on initial test. The others became positive a median of 4 days (IQR 25-75: 3-6) after. Figure 1 illustrates the time-to-event of remaining negative stratified on the presence of symptoms and on the third vaccine dose validity.

**Table 1:** Baseline characteristics of HCWs by RT-PCR result.

| Characteristic | RT-PCR |  | Total |
| --- | --- | --- | --- |
|  | Negative | Positive |  |
| Third vaccine dose valid | 143 | 102 | 245 (51.6%) |
| Absence of symptoms at initial test | 115 | 46 | 161 (33.9%) |
| Initial RT-PCR positive | - | 196 (82.4%) |  |
| Total | 237 | 238 | 475 |

**Table 2:**
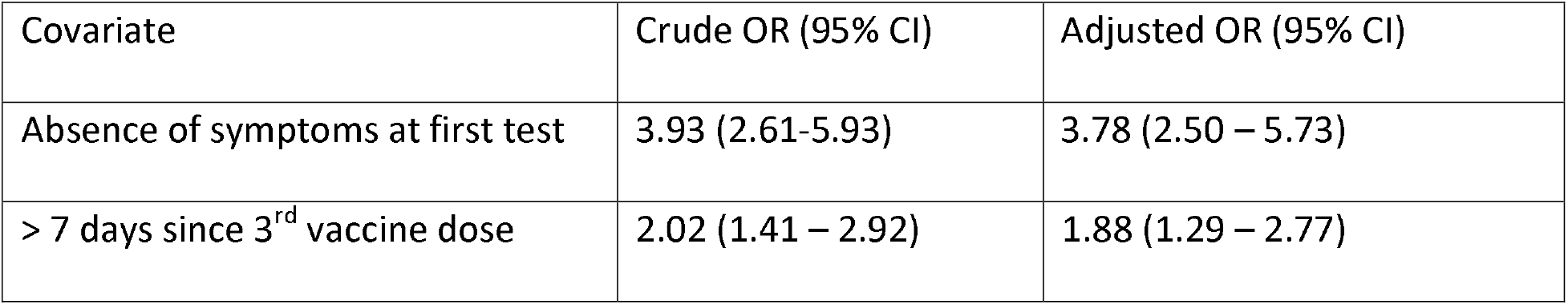
Odds of remaining RT-PCR negative.

**Figure 1:**
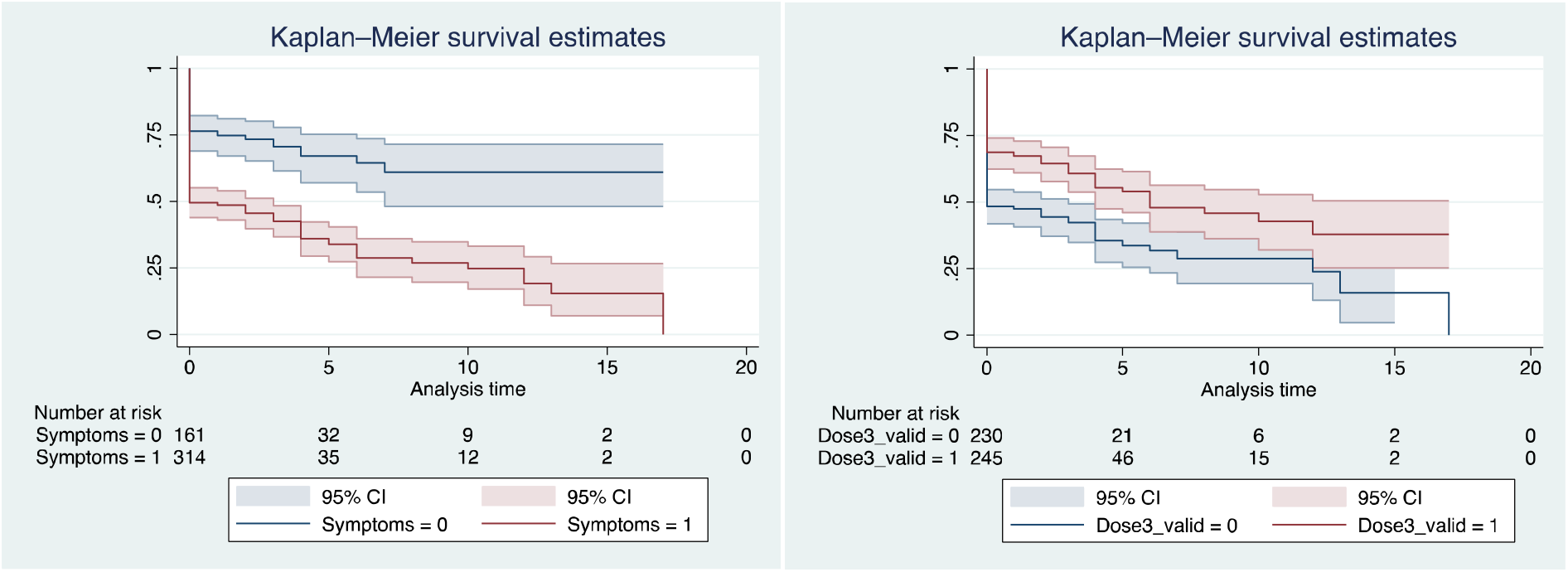
Survival curves for remaining RT-PCR negative (a) stratified by presence of symptoms at first test; (b) stratified by vaccination status.

During that period, a total of 10 outbreaks among HCWs occurred, with a median of 3 HCWs/outbreak (IQR 25-75: 3-6). None were associated with a HCW with a known household contact, although some were associated with a HCW who had a household contact that was identified once contact tracing for the positive HCW was done. There were 9 nosocomial COVID-19 infections identified in patients. In 3 cases, parents and visitors were the source. Three others were exposed to later-declared positive patients in the haematology-oncology day center. For the remaining 3 patients, no source was identified. The list of HCWs who had cared for these 3 patients in the 7 days prior to infection was carefully analyzed and cross-tabulated with HCWs with known household contact who became COVID positive. None were identified as being the source.

## Discussion

We summarized our experience during the SARS-CoV-2 Omicron wave in Montreal (QC), Canada where, for risk-management purposes, our hospital administration opted to allow HCWs with significant household exposures to a confirmed COVID-19 case to work if ≥ 2 doses of vaccines, asymptomatic or with symptoms and a negative PCR result.

Recent studies reported that household secondary attack rates (SAR) were 35.8% in 2021(6) with Omicron having a higher SAR than Delta in a recent study, with boosted individuals had SARs of 25% for Omicron (7). We found a SAR of 50% overall, which decreased to 42% in those boosted. Our SAR is higher than the Danish study, possibly because the outcomes assessment was more complete and done by RT-PCR, while secondary cases in the Danish study could be assessed by rapid antigen detection test, which has a lower sensitivity (5). Moreover, a large proportion of our HCWs were positive upon assessment, questioning the fact that they may not have been a household contact, but either the index case or infected at the same time as their household through a common source.

HCWs that were positive upon initial evaluation were quarantined. Of the remaining 279 HCWs that worked, 42 (15%) became positive. IPAC measures in place mitigated the risk of transmission to patients. The presence of symptoms at initial testing following household exposure was associated with an increased risk of a positive RT-PCR. Adjusting for the presence of symptoms, having a valid 3^rd^ vaccine dose increased the odds of remaining negative by 88%. Our study had some limitations. Although HCWs may not report all known household exposures, they need to be assessed by OHS to have access to their paid sick/contact leaves.

We expect that completeness of data is likely. Given our province-wide availability of RT-PCR results, all results were accessible. As this is real-life setting, it was impossible to have a complete cohort that would include all known and unknown household index cases. Our objective was not to document household SARs but rather to evaluate if it was safe to let HCWs with known household exposures work.

Measures taken to protect the health care environment from COVID-19 must be cautiously balanced with the risk of staffing shortage. Appropriately worn personal protective equipment, is effective against transmission. We thus need to weigh 1) the possible impact of COVID-19 transmission to patients in a mother-child hospital where most would fare well even if infected, 2) the risk of COVID-19 transmission to HCWs whose risk of complications given high vaccination rates is low, to 3) the risk to patients and colleagues (mandatory overtime, errors) of highly specialised staff shortages. Allowing vaccinated asymptomatic HCWs who are known household contacts of confirmed COVID-19 cases to work is likely a safe alternative, when staff shortage is anticipated.

## Data Availability

All data produced in the present study are available upon reasonable request to the authors

## Acknowledgements

This work would not have been possible without the support of the CHU Sainte-Justine teams, including the PCI team: Ms. Gaëlle DeLisle, Ms. Ariane Daoust and Ms. Vicky Gagnon; the nursing call center of the Network Activities Coordination Center and the OHS COVID-19 cell. A special thank you to Marie-Johanne David for her support for the call center and the COVID-19 cell.

CQT is the Tier-1 Canada Research Chair in Infection Prevention and Control: from the Hospital to the Community.

